# A measurement model to quantify systemic bacilli load in severe HIV-associated tuberculosis

**DOI:** 10.1101/2025.07.18.25331763

**Authors:** Bianca Sossen, Charlotte Schutz, Linda Boloko, Tobias Broger, Amy Ward, Abulele Bekiswa, Avuyonke Balfour, Muki Shey, Graeme Meintjes, David Adam Barr

## Abstract

Systemic mycobacterial load is associated with outcome in HIV-associated tuberculosis but is poorly defined. We used latent variable modelling to characterise HIV-associated TB mycobacterial load. *Mycobacterium tuberculosis* detection tests on urine and blood, but not sputum, showed substantial co-variance, and were strongly correlated with host inflammation and mortality.

## INTRODUCTION

Pathogen load is thought to be a major determinant of host response and outcome in infection[1], yet with some exceptions[2–4], the amount of pathogen is generally an unobserved “latent” variable in clinical studies. In tuberculosis, bacilli load has been thought to have a dose-response relationship with host pathology since the rabbit inoculation models of Robert Koch[5]. Classical descriptions of clinical TB based on post mortem pathology held that, in post primary TB (encompassing all extra-pulmonary disease, distinct from cavitatory reactivation pulmonary TB), “the number of bacilli reaching the blood stream and multiplying in the tissues is the main condition” governing the form of disease[6]. Modern post mortem studies of severe HIV-associated TB resonate with these classical studies: fatal HIV-associated TB is disseminated in 90% of cases, as demonstrated by involvement of the reticulo-endothelial system and highly-vascularised organs[7]. Detection of *Mycobacterium tuberculosis* bloodstream infection (MtbBSI) by blood culture is common in severe HIV-associated TB, and is a robust predictor of mortality[8,9]. TB blood culture, however, has imperfect sensitivity[10], and other diagnostics can give additional quantitative information on bacilli load[11].

In summary, systemic bacilli load is considered a fundamental axis of variation in post primary forms of TB and notably in severe HIV-associated TB disease, but cannot be directly measured in clinical studies. Bacilli load may, however, be approximated by multiple, independent TB detection assays that give imperfect but complimentary read-outs in various tissues or samples: such as urine and blood.

The idea of a latent variable of interest reported by multiple observed “indicator” variables is a foundation of many social sciences. Social science approaches - such as classical test theory, latent variable modelling, measurement models, and item response theory - use supervised dimension reduction to summarise covariance in observed variables, quantifying a proposed underlying construct on a tractable scale with reduced measurement error, potentially increasing external validity[12]. Latent variable analysis has also already been used in diagnostic accuracy studies to compensate for imperfect reference standards in extrapulmonary TB[13]. Here, we apply latent variable modeling to TB detection methods to define systemic or MtbBSI bacilli load in patients with severe HIV-associated TB.

## METHODS

### Study Design, Patients and Procedures

This analysis was performed on data from a previously reported large, prospective observational cohort study which recruited participants admitted to a district hospital in Khayelitsha, Cape Town, with a new diagnosis of HIV-associated TB[8]. In brief, participants were adults with CD4 counts <350cells/μl, and had sputum (with induction if needed), urine and blood samples collected prospectively for TB testing and storage of neat urine and whole blood. Enrolment occurred within a median of two days of presentation to hospital and samples were collected on this enrolment day or the day thereafter. Only participants with microbiologically confirmed TB (including via Xpert MTB/RIF (Cepheid, USA), mycobacterial culture or urine Determine^TM^ Alere Antigen (AlereLAM; Abbott, USA)) were included in the present analysis. For the purposes of this analysis, vital status outcome at 28 days was assessed.

When sputum was obtained, it was sent in real time for mycobacterial culture and Xpert MTB/RIF (Sputum-XP) at the centralized, accredited South African National Health Laboratory Service (NHLS). Mycobacterial blood cultures were performed in real time on whole blood in Myco/F-Lytic (Becton Dickinson Biosciences) bottles at NHLS. On any sputum or blood cultures that were positive, the time-to-positivity (TTP) was recorded in number of days and the GenoType MTBDR*plus* assay (Hain Lifesciences) was used to identify Mtb. When urine was obtained, it was stored at -80°C for LAM testing and the remainder was concentrated for urine Xpert MTB/RIF (Urine-XP) testing in real time. Frozen urine was defrosted towards SILVAMP TB LAM (FujiLAM; Fujifilm, Japan) and AlereLAM testing in 2018[14]. For AlereLAM, 60μl urine was applied to the sample pad and the result read as per the manufacturer’s reference card at 25 minutes. FujiLAM was tested according to manufacturer instructions. Any visible FujiLAM patient line was considered positive. To obtain optical density (OD) quantitative values of the patient line, a Western Blot reader was used for both LAM assays. Those with stored whole blood available had blood Xpert ultra (Blood-XPU) tested as previously reported[11]. On all samples, the Xpert MTB/RIF Ultra provides an automated result of positive or negative for Mtb and gives a quantitative result of cycle threshold (Ct) value. The study was approved by the University of Cape Town Human Research Ethics Committee, as previously reported[8].

### Statistical analysis

The included TB detection assays (Sputum-XP, Urine-XP, Blood-XPU, FujiLAM, AlereLAM, blood and sputum culture) generate a semi-quantitative read-out in case of a positive result (TTP, Ct value or OD). Each was converted to a 4-level ordinal scale where negative results were assigned as “0”, and the range of semi-quantitative read-outs divided into three intervals, from “1” (highest Ct/TTP, lowest OD) to “3” (lowest Ct/TTP, highest OD) interval. Missing assay results were imputed using multivariate imputation by chained equations.

Ordinal scale polychoric correlations between assays were calculated and patterns of covariance assessed using hierarchical clustering and exploratory factor analysis. Evidence for number of dimensions was assessed using scree plotting, parallel analysis, and Bayesian Information Criteria (BIC). Extracted dimensions were assessed for relationship to clinical phenotype using network graphing. A graded response model (GRM) was fit using the R Multidimensional Item Response Theory (IRT) package (v1.43)[15]. GRM is applicable when “item responses” (in this case, TB assay results) are reported on an ordered polytomous scale. In GRM, a latent variable (“*theta*”) is related to observed item results by a logistic curve with a slope parameter and thresholds that identify boundaries on an ordered outcome - analogous to an ordinal logistic regression, where the predictor variable is unobserved but estimated by the model. Items (in this case TB detection assays) with good discrimination for *theta* (better signal to noise) have higher 2-slope parameters, but items can also vary in the threshold of *theta* at which they (probabilistically) move from one ordinal category to the next. Items with poorer discrimination may therefore still give information at levels of *theta* not well reported on by higher discrimination items, and an advantage of IRT models is an ability to characterise individual item performance.

## RESULTS

Data from 519 hospitalised participants with confirmed HIV-associated tuberculosis were included. Participants had a median age of 36 years, median CD4 of 55 cells/mm^3^ and 50.7% were female (supplementary table 1). Of the seven TB detection assays in 519 individuals, 85% of assay results were available; most missing results were sputum culture and Xpert, from non-production of a sputum sample (supplementary table 1). The distribution of semi-quantitative read-outs from the seven assays (OD, TTP, and Ct) were, in general, skewed and with evidence of ceiling and floor effects (figure 1A). Assays showed collinearity by sample compartment: strongest correlations were blood culture with blood Xpert, and sputum culture with sputum Xpert (figure 1B&C). Urine assays were also significantly collinear, but urine Xpert was more closely related to blood Xpert than urine LAM positivity (figure 1C).

**Figure 1:**
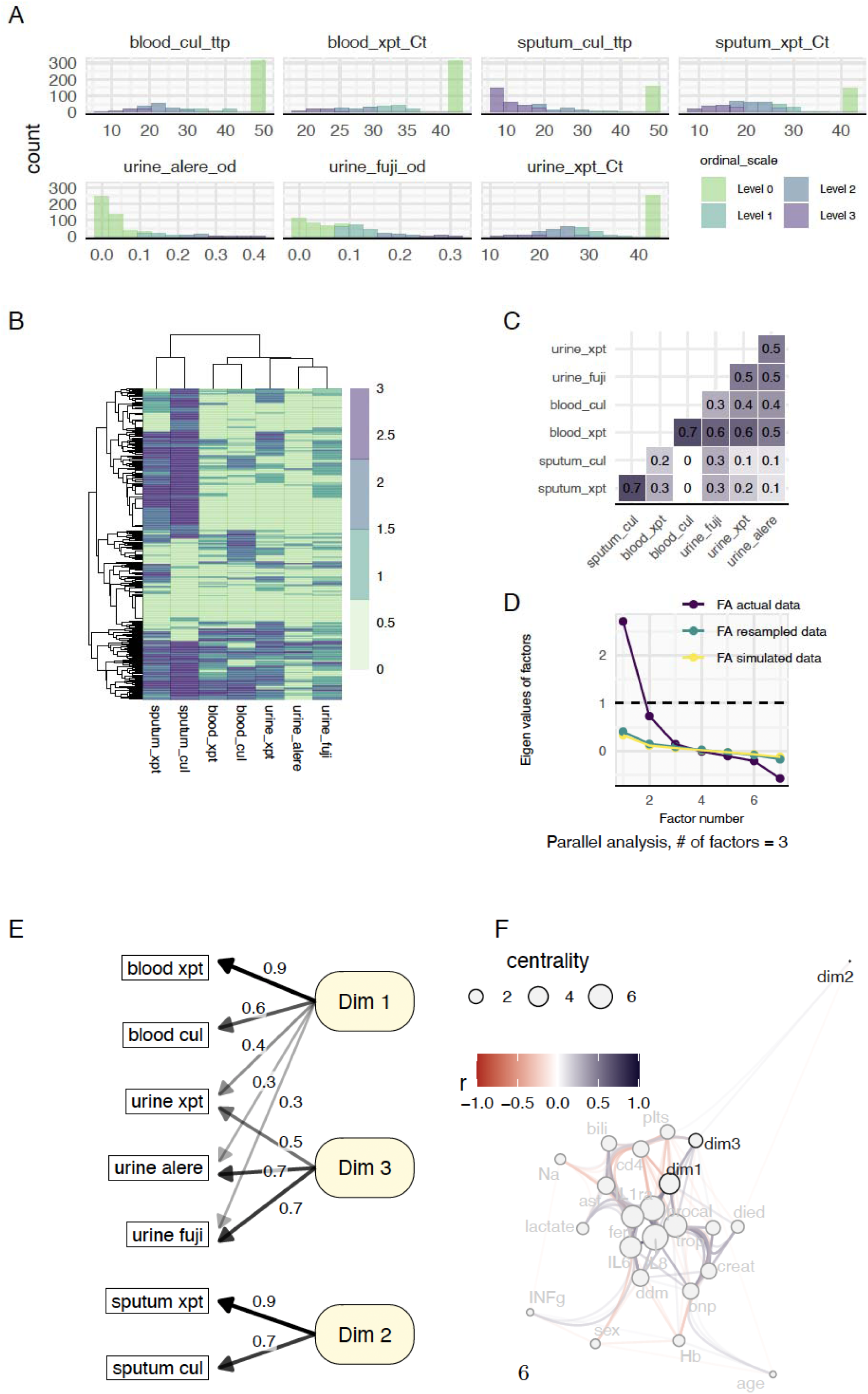
Co-variance structure in TB detection assay variables measured on polytomous ordinal scale reveals three latent bacilli load variables. A. Histograms of Culture Time to Positivity (ttp), Xpert PCR Cycle Threshold (Ct), and urine LAM Optical Density (od), with below limit of detection values shown at limit of detection for culture and Xpert. Histogram bars are coloured by polytomous ordinal scale category 0-3 used in subsequent analysis. For negative Xpert and culture samples, level 0 is assigned to negative tests (below limit of detection). B. Hierarchically clustered heat map of the seven TB assay results on ordinal scale (columns) for each participant (rows). Mminkowski distance measure and complete linkage used. C. Polychoric Correlation matrix for the seven assays’ polytomous ordinal scale results. D. Scree plot and parallel analysis on polychoric correlation matrix, used to determine number of factors, i.e. dimensions of covariance, in the seven TB assay results. Factor analysis is shown for original data with eigen values for extracted factors (purple line); the same is shown for re-sampled and simulated (random) versions of the data: three factors in data contain more information (higher eigen values) than the resampled or simulated versions of the data suggesting three factors contain signal above that expected for random noise. Three factor exploratory factor model also has lower BIC than a two-factor model (BIC 31 versus -15 respectively) also supporting the presence of 3 rather than 2 dimensions in the data. E. Factor loadings (shown by numbers on arrows) for a three-factor model applied to the seven TB test data. Only the statistically significant loadings are shown. The first factor (Dim 1) loads most strongly on blood Xpert ultra (blood Xpt) but also loads blood culture (blood cul) and urine Xpert (urine xpt) and weakly loads the two urine LAM tests (Alere and Fuji). Dim 3 loads covariance in urine tests not captured on Dim 1. The sputum variables (culture and Xpert) load separately in their own factor (Dim 2). F. Network analysis graph showing correlation structure for 20 clinical meta-data variables plus varimax rotated factor scores for the three factor TB test factor analysis model (Dim 1, Dim 2 and Dim 3). Edge transparency and colour show strength and direction of correlation respectively. Node (vertex) size shows centrality of the variable in the correlation network, calculated from node degree (number of connections) weighted by absolute correlation strength. Nodes are placed according to the force-directed algorithm of Fruchterman and Reingold, in which closely connected nodes are represented near each other but without overlapping. Edges (connections) are bundled by letting edges follow the shortest path along the graph’s minimal spanning tree. This demonstrates that Dim 1 (blood and urine test covariance) is a variable with high centrality (strong correlation with many clinical meta-data variables); by comparison, Dim 3 (urine test results covariance unrelated to Dim 1) is more peripheral, while Dim 2 (sputum test results) is almost completely unrelated to other clinical variation. lactate = blood Lactate; Na = serum sodium; died = died by day 28 follow up; IL1ra = interleukin 1 receptor agonist in plasma; trop = high sensitivity troponin t in serum; bnp = N-terminal pro b-type natriuretic peptide in serum; IL8 = interleukin 8 in plasma; IL6 = interleukin 6 in plasma; Hb = peripheral blood haemoglobin; creat = serum creatinine; INFg = interferon gamma in plasma; sex = 1 if female; age = participant age in years; bili = total serum bilirubin; ast = serum aspartate aminotransferase; ddm = blood d-dimer; ferr = serum ferritin; cd4 = peripheral blood CD4 cell count.

Parallel analysis suggested that covariance in the seven assays could be summarised in three dimensions (figure 1D). In a three-factor model, the first dimension predominantly represented blood Xpert and blood culture variance, but also with significant loading of urine Xpert and urine LAM (figure 1E). Dimension 2 exclusively loaded variation in sputum assays, and dimension 3 loaded residual variation in urine LAM and urine Xpert that was not captured on dimension 1 (figure 1E).

Next, we assessed how closely related each of the three TB assay factors were to important clinical variables. Dimension 1 (blood and urine assay variance) was highly correlated with multiple host response variables, showing high centrality in a network correlation graph (figure 1F). Dimension 3 (residual urine assay variance) was more peripheral in the clinical variable network, while dimension 2 (sputum assays) was almost completely unrelated to any clinical variables (figure 1F).

We postulated that dimension 1 was most in keeping with a read-out of systemic bacilli load (while dimensions 2 and 3 were summarising variance specific to sputum and urine compartments). Sputum assays were dropped from further analysis and a Graded Response Model (GRM) comprising the 5 remaining (blood and urine) assays was fit as a measurement model for systemic bacilli load. This replicated dimension 1 from the previous factor analysis, estimating a latent variable, *theta*, which was most strongly related to blood Xpert ultra and blood culture variance, but again also loading covariance of the three urine assays (figure 2 A&B).

**Figure 2.**
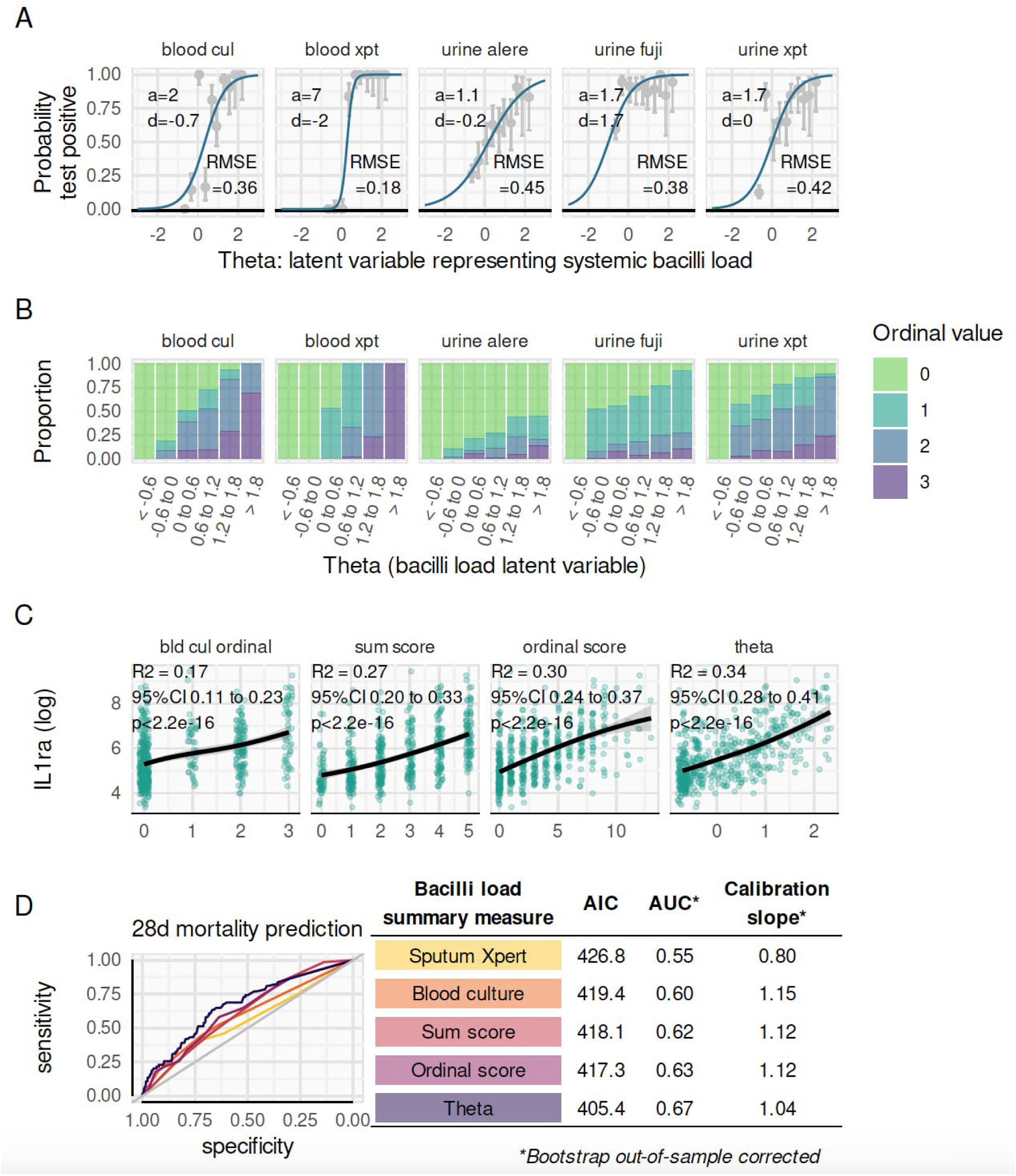
Graded response model for measuring systemic bacilli load. A. A two factor graded response model was fit to the ordinal scale versions of TB blood culture, blood Xpert ultra, urine AlereLAM, urine FujiLAM, and urine Xpert results using the mirt() package. Varimax rotated factor scores corresponding to dimension 1 from figure 1 were extracted as a summary measure, theta, for the underlying latent variable of interest i.e. systemic bacilli load. Shown are two parameter logistic models [of form y = 1/(1 + exp( -(a °ø theta + d) ))] relating this latent variable (theta) to test positivity (y, moving from 0 to 1 on ordinal scale) for the five TB detection assays. Estimates for the slope (a) and location/threshold (d) parameters for each test are given and show that, from the five TB tests, blood Xpert ultra has the steepest slope (highest discrimination) value for theta. Model fit is shown with overlaid proportion of positive tests within each decile of theta (grey points with 95% confidence intervals), and summarised with root-mean-squared-error (RMSE) values; these show that theta is more closely related to blood Xpert ultra positivity, followed by blood culture and then urine tests. B. Ordinal level of TB detection assays by level of theta. As theta rises, all TB assays have an increased probability of higher ordinal category. Blood Xpert ultra has highest discrimination for theta, moving rapidly from 0 (below limit of detection) to 3 (highest ordinal category): all participants with theta > 1.8 have blood Xpert value of 3. However, at low values of theta (< 0) blood Xpert ultra gives limited information as this assay is uniformly below LoD. Other assays, such as urine FujiLAM give more granular results at low levels of theta despite having poorer discrimination overall. C. Comparative strength of association between three proposed summary measures of systemic bacilli load and plasma interleukin 1 receptor agonist, IL1ra, on log scale. The proposed summary measures of systemic bacilli load are an ordinal version of blood culture TTP; a sum score, which sums the total number of tests positive from TB blood culture, blood Xpert ultra, urine AlereLAM, urine FujiLAM, and urine Xpert; ordinal score which sums ordinal scale versions of these five TB tests (thus capturing semi-quantitative read-outs from these tests); and *theta* (the latent variable factor scores) from the graded response model in A. IL-1ra was chosen (a priori) to represent a host response variable hypothesized to be an important mediator of pathophysiology in prior work. Of the three bacilli load summary measures, *theta* has strongest association with host response patho-phenotype, ‘explaining’ 34% of variance in log IL1ra compared to 17%, 27% and 30% for blood culture ordinal scale, sum score and ordinal score respectively. D. Theta is also more strongly associated with mortality than the other summary measures of bacilli load: sputum Xpert Ct ordinal scale, blood culture TTP ordinal scale, sum score and ordinal score. Discrimination for day 28 mortality (bootstrapped ROC AUC) is higher at 0.67 compared to 0.55, 0.60, 0.62, and 0.63 for logistic regression models relating theta, sputum Xpert Ct ordinal scale, blood culture TTP ordinal scale, sum score, or ordinal score, to day 28 mortality respectively. Mortality prediction calibration (assessed by bootstrapped calibration slope closer to 1) and overall model fit (assessed by lower Akaike information criterion value) is also better for theta than the other measures

We compared *theta* from the GRM model to three other measures of systemic or blood bacilli load: blood culture positivity and TTP measured on an ordinal scale; a simple sum score, totaling the number of positive blood and urine assays (range 0 to 5), an approach used previously[8,15]; and an ordinal score summing the ordinal category values across the 5 assays (range 0 to 15). Sum score, ordinal score and *theta* were all highly correlated (Rho > 0.88 for all 3 pairwise comparisons). Correlation with IL-1-receptor antagonist (IL-1ra, chosen as most central node in the correlation network graph, figure 1F, and because of prior strong association with mortality [8]) varied (figure 2C). Blood culture positivity and TTP explained the least variance in IL-1ra (17%) while theta explained the most (34%). Similarly, theta had the greatest predictive accuracy (highest AUC, best calibration, and lowest AIC) for day 28 mortality (figure 2D).

## DISCUSSION

Disseminated TB, while poorly defined, is a severe form of TB that has a high risk of rapid clinical deterioration and death[16]. Disseminated TB definitions, such as that from the World Health Organisation of “detection of Mycobacterium tuberculosis (MTB) from more than one clinical site”[17] are subject to substantial ascertainment bias and are unlikely to be reproducible. Bacteraemic and non-bacteraemic disseminated TB disease may be clinically distinct entities[18], and classical descriptions of post-primary TB place major emphasis on bacilli load in blood[6,18].

We found that TB detection assays show patterns of covariance in keeping with underlying systemic bacilli load. We formally defined this bacilli load latent variable using a measurement model: incorporating urine and blood PCR Ct values, urine LAM antigen levels, and blood culture TTP. Previously, we have used sum scores of TB detection assays in blood and urine to define bacilli load[8,16]. The latent variable measurement approach described here outperformed sum scoring, as evidenced by increased variance explained in the host response biomarker IL-1ra from 27% to 34%, and improved discrimination and calibration for mortality prediction. This impact on effect size of association with important clinical variables, external to the measurement model, can be interpreted as strong evidence of reduced measurement error and increased external validity.

Previously, microbial load has been associated with pathophysiology and outcome in clinical infections where reliable quantitative pathogen read-outs are available. For example, concentration of lipopolysaccharide determines disease phenotype, cytokine release levels and mortality risk in *Neisseria meningitidis* infection,[4] while *Plasmodium falciparum* quantitative parasitaemia is a major predictor of mortality and explains most variance in host gene expression.[18] By contrast, TTP of blood cultures in pyogenic bacteraemia has shown conflicting correlation with mortality.[19] We found mycobacterial blood culture positivity and TTP to be a relatively weak predictor, but show that combining multiple alternative read-outs (including weak predictors like blood culture TTP) using latent variable modelling, resulted in a pathogen load read-out which related strongly to mortality and host response. This suggests that weak association of blood culture TTP does not mean that (the latent variable) pathogen load is unimportant, but rather that TTP is an indicator variable biased towards the null by measurement error.

A limitation of this study is that some of the specific TB tests used in the analysis are not widely available (in particular, the first generation FujiLAM assay is no longer manufactured) which precludes direct validation in future studies. However, one advantage of the item response theory model – in contrast to, for example, sum scores – is that estimates of the underlying latent variable is still possible when an individual item is not observed (albeit with added uncertainty).

Measurement model approaches may be a useful method to define latent variables, such as pathogen load, in clinical studies. The ability to quantify systemic, or bloodstream, bacilli load could lead to a better definition of disseminated TB, facilitating improved research into this severe condition.

## Data Availability

Data from the present study are available upon reasonable request to the corresponding author

## SUPPLEMENTARY

**Supplementary table 1.**
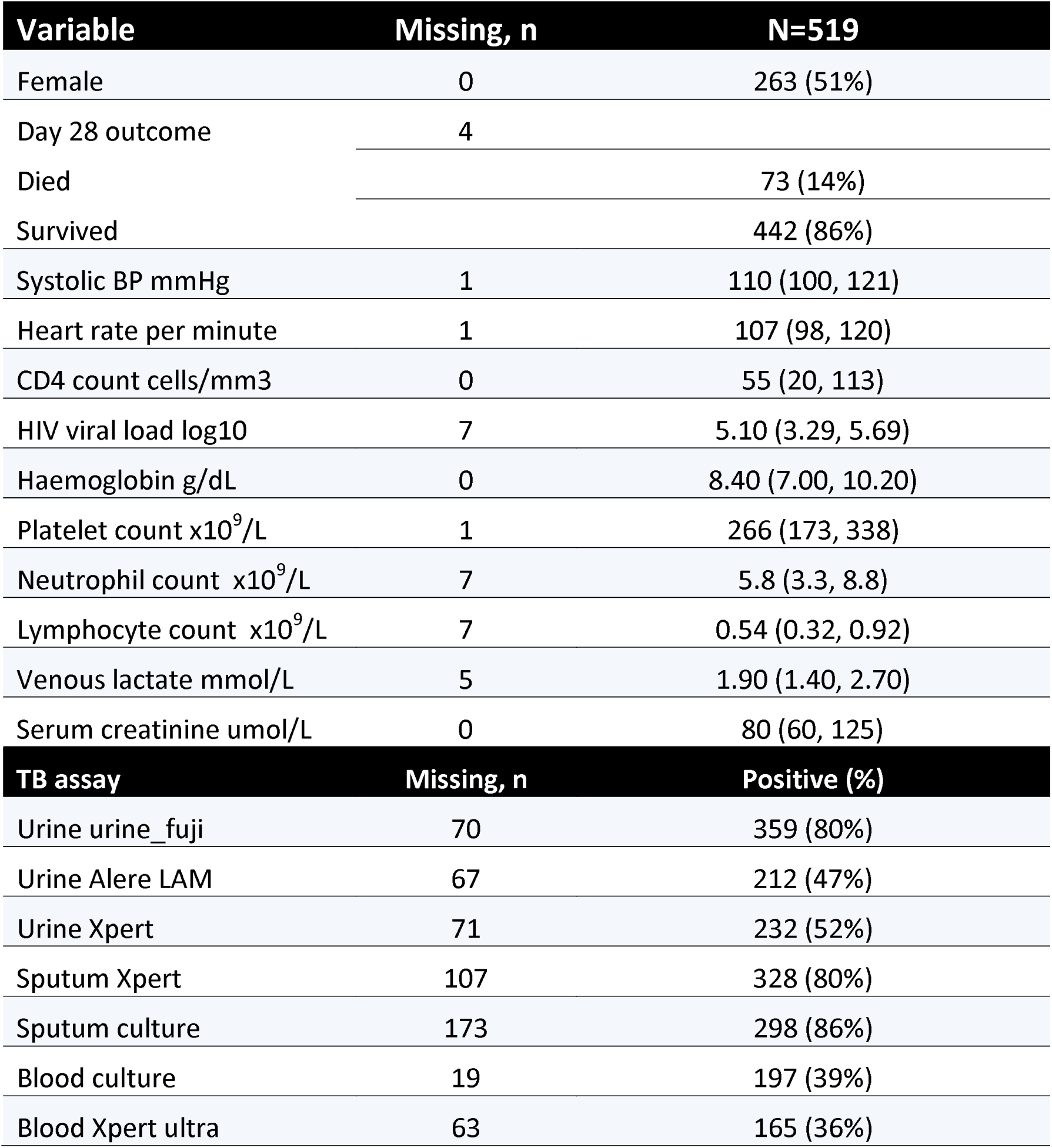

| Variable | Missing, n | N=519 |
| --- | --- | --- |
| Female | 0 | 263 (51%) |
| Day 28 outcome | 4 |  |
| Died |  | 73 (14%) |
| Survived |  | 442 (86%) |
| Systolic BP mmHg | 1 | 110 (100, 121) |
| Heart rate per minute | 1 | 107 (98, 120) |
| CD4 count cells/mm3 | 0 | 55 (20, 113) |
| HIV viral load log10 | 7 | 5.10 (3.29, 5.69) |
| Haemoglobin g/dL | 0 | 8.40 (7.00, 10.20) |
| Platelet count x10<sup>9</sup>/L | 1 | 266 (173, 338) |
| Neutrophil count x10<sup>9</sup>/L | 7 | 5.8 (3.3, 8.8) |
| Lymphocyte count x10<sup>9</sup>/L | 7 | 0.54 (0.32, 0.92) |
| Venous lactate mmol/L | 5 | 1.90 (1.40, 2.70) |
| Serum creatinine umol/L | 0 | 80 (60, 125) |
| TB assay | Missing, n | Positive (%) |
| Urine urine_fuji | 70 | 359 (80%) |
| Urine Alere LAM | 67 | 212 (47%) |
| Urine Xpert | 71 | 232 (52%) |
| Sputum Xpert | 107 | 328 (80%) |
| Sputum culture | 173 | 298 (86%) |
| Blood culture | 19 | 197 (39%) |
| Blood Xpert ultra | 63 | 165 (36%) |

## REFERENCES

1. Wiersinga WJ, Van Der Poll T. Immunopathophysiology of human sepsis. eBioMedicine 2022; 86:104363.

2. Towner JS, Rollin PE, Bausch DG, et al. Rapid Diagnosis of Ebola Hemorrhagic Fever by Reverse Transcription-PCR in an Outbreak Setting and Assessment of Patient Viral Load as a Predictor of Outcome. J Virol 2004; 78:4330–4341.

3. Fajnzylber J, Regan J, Coxen K, et al. SARS-CoV-2 viral load is associated with increased disease severity and mortality. Nat Commun 2020; 11:5493.

4. Møller AW, Bjerre A, Brusletto B, Joø GB, Brandtzaeg P, Kierulf P. Chemokine Patterns in Meningococcal Disease. J Infect Dis 2005; 191:768–775.

5. Carter K. Essays of Robert Koch. Praeger, 1987. Available at: https://www.bloomsburycollections.com/monograph?docid=b-9798400647277.

6. Pagel W. An Outline of the Principal Forms of Tuberculosis in Man. Postgrad Med J 1952; 28:606–614.

7. Gupta RK, Lucas SB, Fielding KL, Lawn SD. Prevalence of tuberculosis in post-mortem studies of HIV-infected adults and children in resource-limited settings: a systematic review and meta-analysis. AIDS 2015; 29:1987–2002.

8. Schutz C, Barr D, Andrade BB, et al. Clinical, microbiologic, and immunologic determinants of mortality in hospitalized patients with HIV-associated tuberculosis: A prospective cohort study. PLOS Med 2019; 16:e1002840.

9. Barr DA, Lewis JM, Feasey N, et al. Mycobacterium tuberculosis bloodstream infection prevalence, diagnosis, and mortality risk in seriously ill adults with HIV: a systematic review and meta-analysis of individual patient data. Lancet Infect Dis 2020; 20:742–752.

10. Barr DA, Kerkhoff AD, Schutz C, et al. HIV-Associated *Mycobacterium tuberculosis* Bloodstream Infection Is Underdiagnosed by Single Blood Culture. J Clin Microbiol 2018; 56:e01914–17, /jcm/56/5/e01914-17.atom.

11. Boloko L, Schutz C, Sibiya N, et al. Xpert Ultra testing of blood in severe HIV-associated tuberculosis to detect and measure Mycobacterium tuberculosis blood stream infection: a diagnostic and disease biomarker cohort study. Lancet Microbe 2022; 3:e521–e532.

12. Brown TA. Latent Variable Measurement Models, in Todd D Little (ed.), The Oxford Handbook of Quantitative Methods in Psychology: Vol. 2: Statistical Analysis. Oxford Library of Psycohology, 2013. Available at: 10.1093/oxfordhb/9780199934898.013.0013. Accessed 21 February 2025.

13. MacLean EL, Kohli M, Köppel L, et al. Bayesian latent class analysis produced diagnostic accuracy estimates that were more interpretable than composite reference standards for extrapulmonary tuberculosis tests. Diagn Progn Res 2022; 6:11.

14. Broger T, Sossen B, du Toit E, et al. Novel lipoarabinomannan point-of-care tuberculosis test for people with HIV: a diagnostic accuracy study. Lancet Infect Dis 2019; 19:852–861.

15. Chalmers RP. mirt: A Multidimensional Item Response Theory Package for the R Environment. J Stat Softw 2012; 48. Available at: http://www.jstatsoft.org/v48/i06/. Accessed 24 January 2025.

16. Kerkhoff AD, Barr DA, Schutz C, et al. Disseminated tuberculosis among hospitalised HIV patients in South Africa: a common condition that can be rapidly diagnosed using urine-based assays. Sci Rep 2017; 7:10931.

17. Providing Care to People with Advanced HIV Disease Who Are Seriously Ill Policy Brief. Geneva: World Health Organization, 2023.

18. Crump JA, Reller LB. Two Decades of Disseminated Tuberculosis at a University Medical Center: The Expanding Role of Mycobacterial Blood Culture. Clin Infect Dis 2003; 37:1037–1043.

